# The relationship between hyperglycaemia on admission and patient outcome is modified by hyperlactatemia and diabetic status: a retrospective analysis of the eICU collaborative research database

**DOI:** 10.1101/2023.05.01.23289339

**Authors:** Oisin Fitzgerald, Oscar Perez-Concha, Blanca Gallego-Luxan, Lachlan Rudd, Louisa Jorm

## Abstract

Both blood glucose and lactate are well-known predictors of organ dysfunction and mortality in critically ill patients. Previous research has shown that concurrent adjustment for glucose and lactate modifies the relationship between these variables and patient outcomes, including blunting of the association between blood glucose and patient outcome. We aim to investigate the relationship between ICU admission blood glucose and hospital mortality while accounting for lactate and diabetic status. Across 43,250 ICU admissions, weighted to account for missing data, we assessed the predictive ability of several logistic regression and generalised additive models that included blood glucose, blood lactate and diabetic status. We found that inclusion of blood glucose marginally improved predictive performance in all patients: AUC-ROC 0.665 vs 0.659, with a greater degree of improvement seen in non-diabetics: AUC-ROC 0.675 vs 0.663. Inspection of the estimated risk profiles revealed the standard U-shaped risk profile for blood glucose was only present in non-diabetic patients after controlling for blood lactate levels. Future research should aim to utilise observational data to estimate whether interventions such as insulin further modify this effect, with the goal of informing future RCTs of interventions targeting glycaemic control in the ICU.

## Introduction

Both blood glucose and lactate are well-known predictors of organ dysfunction and mortality in critically ill patients [1, 2]. Glucose shows a U-shaped relationship with mortality with both hypo- and hyperglycaemia associated with poor outcomes. The association between hospital mortality and lactate is particularly strong, with previous research demonstrating lactate has comparable predictive ability to the APACHE-II, SOFA and qSOFA scores in certain ICU populations [3]. Further, it has been shown in previous research that concurrent adjustment for glucose and lactate modifies the relationship between these variables and patient outcomes, including blunting of the association between blood glucose and patient outcome [3–6].

The metabolic pathways of glucose and lactate are highly inter-connected. Lactate is an end-product of glycolysis, the oxygen independent cellular respiration of glucose, and is a major substrate for gluconeogenesis in the liver and kidney [7]. In previous work, our team has found that blood lactate is the most important non-glucose blood gas/laboratory factor in predicting future blood glucose in critically ill patients [8]. The mechanisms that lead to hyperglycaemia (>180 mg/dL) and hyperlactatemia (>2 mmol/L) in critical illness are complex and varied, with clinical interventions playing a role [9–11]. While tissue hypoxia leading to increased anaerobic respiration has commonly been seen as the primary cause of increased lactate levels, this view has been challenged, with both hyperlactatemia and hyperglycaemia also linked to metabolic alterations associated with immune activation [7, 12–14]. A major question for clinicians and researchers is when components of this response are maladaptive requiring treatment.

The degree to which blood glucose (at varying thresholds) is a marker or mediator of poor patient outcomes has been debated [15, 16], with observational studies demonstrating lack of, or attenuated, association between blood glucose and patient outcome in certain circumstances [3–6], and randomised control trials (RCTs) demonstrating that tight glycaemic control (a target 80-110 mg/dL) is associated with poorer outcomes than a less stringent target of <180 mg/dL [17]. The pathway from short term glucose toxicity to poor outcomes in the critically ill has not been fully elucidated, with organ dysfunction as a result of glucose induced inflammation and oxidative damage one hypothesised route [18, 19]. However, elevated lactate may also have deleterious effects, for instance through immunosuppression [14, 20], raising the potential that elevated blood glucose levels may be a marker of this effect.

Accordingly, we aim to investigate whether blood glucose is an independent predictor of hospital outcome while controlling for blood lactate across subgroups defined by diabetic status. To account for non-linear relationships, we use flexible semi-parametric statistical models which retain the benefit of being readily interpretable while accounting for non-linear effects [21].

## Methods

### Patients and data sources

Data for this study were sourced from the eICU collaborative research database (eICU-CRD) open access critical care database, de-identified to conform with the Health Insurance Portability and Accountability Act (HIPAA). eICU-CRD is a large multi-center critical care database holding data associated with 200,859 ICU stays admitted at 208 hospitals across the United States between 2014 and 2015 [22]. As described in previous research [23, 24] data from the eICU-CRD are generally of high quality, with common vital signs (such as blood glucose) and patient outcomes well recorded, but missing data creating challenges identifying which patients received complex interventions such as intubation, ventilation, and dialysis beyond the first day of ICU stay and in the calculation of risk scores that depend on non- bedside measurements Institutional ethics for the research project was obtained from UNSW Sydney - HC220829.

### Inclusion criteria and data quality assessment

We restrict our analysis to non-elective adult patients (over 18-years-old) who had an ICU stay more than 12 hours in duration, received an APACHE-IV score and were not admitted for diabetic ketoacidosis or hyperosmolar hyperglycaemic state. From this cohort we selected all patients who had at least one glucose and lactate measurement within -12 to +24 hours of their ICU admission (see Figure 1).

**Figure 1.**
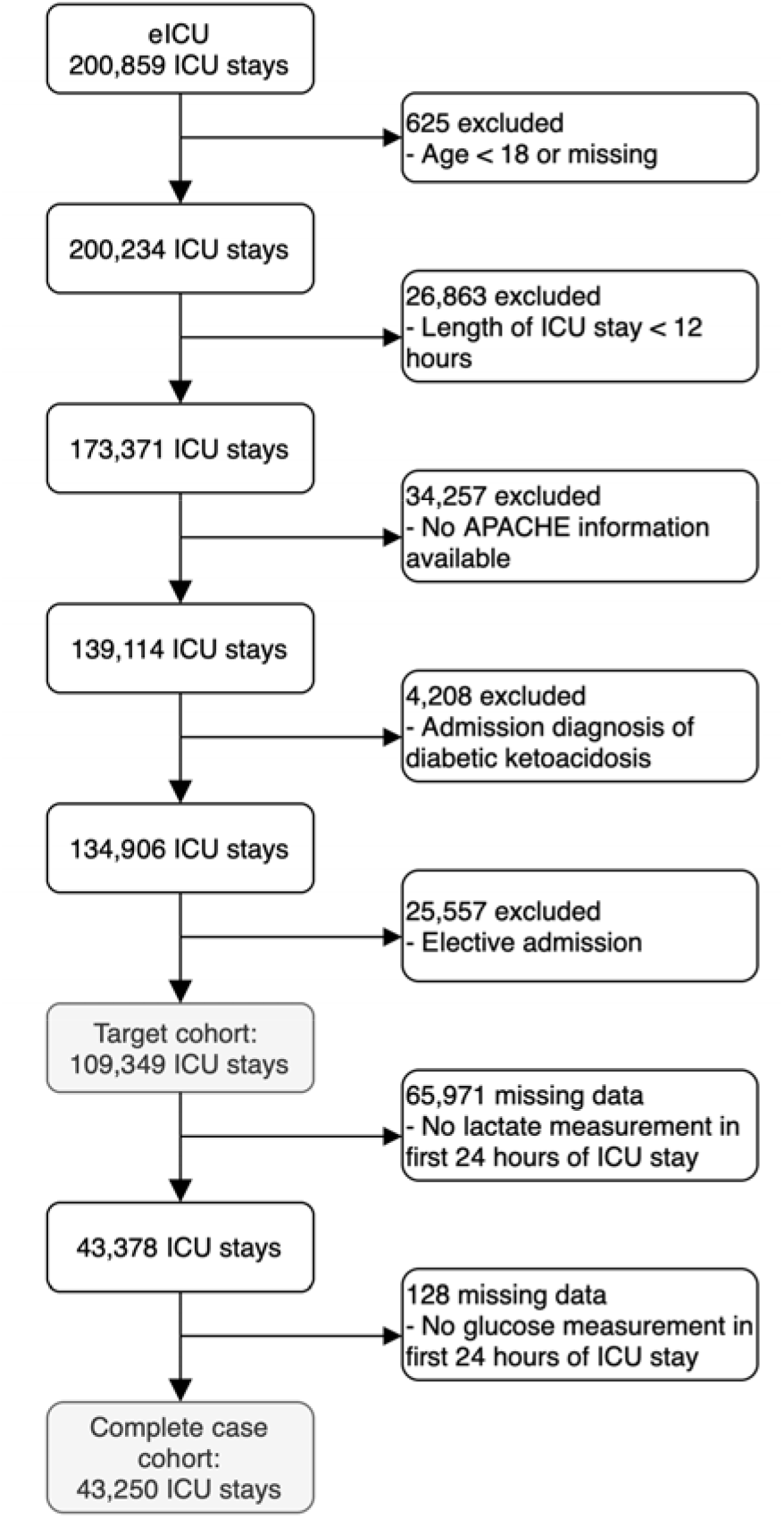
Impact of the inclusion criteria and missing data on extraction of the target and final complete case cohort from the eICU-CRD database.

### Data extraction

All data extraction and transformation processes were carried out using the R packages bigrquery [25] and data.table [26]. All queries are freely available at the project code repository (linked below). The following variables on admission (-12 to +24 hours of index admission time) were extracted from the database: ICU type, age, gender, ethnicity, weight, BMI, diabetic status, Elixhauser comorbidity index, APACHE admission diagnosis, APACHE-IV score, lactate, glucose, bilirubin, potassium, sodium, chloride, creatinine, blood urea nitrogen, calcium, along with indication of the use or prescription of insulin, intubation and ventilation. Extracted outcomes included in-hospital and in-ICU mortality and length of ICU stay.

### Data availability

The data underlying this article are freely available at https://eicu-crd.mit.edu/ and can be accessed following completion of the required training and data usage agreements.

### Statistical analysis

#### Matching blood glucose and lactate measurements

As blood lactate is measured less frequently than blood glucose, we construct our blood glucose and lactate variables by: 1) selecting the blood lactate measurement closest to ICU admission and 2) taking the average of all blood glucose measurements within 1-12 hours of this blood lactate measurement. The size of the blood glucose averaging window is the minimum value (at 1 hour increments) from 1-12 such that at least one blood glucose measurement is available.

#### Descriptive statistics

We give a descriptive analysis of the data using graphs of the key variables and through summarising the dataset in a table. Graphically we report the univariate and bivariate relationship between blood glucose and blood lactate, stratified by diabetic status and hospital mortality status. To avoid overplotting we show the bivariate relationships using contour plots for the stratified results. In the tables, continuous variables are reported using the mean and standard deviation. Categorical variables are reported as percentages. Data are reported stratified by whether a lactate measurement was available.

#### Statistical modelling of missingness

We use inverse probability weighting to account for potential selection biases introduced by the irregular measurement of blood lactate [27]. This approach uses the complete cases to create a pseudo-population designed to mimic the characteristics of the target population (in this case adults with an ICU stay over 12 hours and an APACHE-IVa score [28]). We use a logistic regression model to calculate the probability that blood lactate would be measured in our target population using the following covariates: age, APACHE-IVa score, operative admission, an APACHE admission diagnosis of sepsis, APACHE admission organ system and ICU type. We assess the model fit using the area under the ROC curve (AUC-ROC) and a calibration plot. For each row in the complete case dataset the weights are calculate as the inverse of the model prediction.

We performed several sensitivity analyses to ensure our results are not overly impacted by our approach to account for missing data. We use a machine learning model (XGBoost [29]) to additionally calculate the missingness weights using the same set of variables as the logistic regression model, along with all non-lactate laboratory results (see Data Extraction above). To avoid overfitting bias this was done using cross-validated out of sample prediction [30]. Additionally, we develop a linear generalised additive model (GAM) and XGBoost imputation models, that were fit on the subsample of observations with a lactate measurement. These use the same variables sets and methodologies (i.e., out of sample prediction for the XGBoost model) as the missingness models, and in the imputation analysis their prediction replace the unmeasured lactate values for the 65,971 observations with no recorded blood lactate.

#### Statistical modelling of hospital mortality

The aim of the statistical modelling of hospital mortality was: 1) to assess the degree to which blood glucose is predictive of mortality when controlling for blood lactate and diabetic status (model selection step) and 2) describe the functional form of this relationship (model interpretation step).

##### Models

We use logistic and GAMs to build the models. GAMs are additive models that allow the predictor effects to vary non-linearly with the level of the covariate by transforming the variable through some smooth function *f*, e.g., a spline parameterisation. For a given basis dimension, GAMs incorporate the penalization of the spline and model estimation into a single process reducing the need for model hyperparameter tuning compared to pre model fitting spline transformation of a regression variable [21]. We use thin-plate regression splines [31]. In cases where the smooth spline is a two-dimensional interaction effect we denote this by *var1 : var2*. For the logistic regression (LR) models, we transformed glucose and lactate into categorical (binned) variables using the cut points from Freire Jorge, Wieringa [6]. We fit the following models, for all observations and stratified by diabetic status, using the iterative fitting method introduced in Wood [21] for GAMs and maximum likelihood for logistic regression models:

1. LR: glucose
2. LR: lactate
3. LR: glucose + lactate
4. LR: glucose + lactate + glucose : lactate
5. GAM: glucose
6. GAM: lactate
7. GAM: glucose + lactate
8. GAM: (glucose : insulin)

##### Model selection

Mirroring the aims, the statistical analysis was performed in two steps: model selection and model interpretation. During the model selection step a weighted 10- fold cross-validation (CV) procedure was used to assess model fit [32]. The splits were made using the patient health care system ID, ensuring that all ICU stays from a patient were either in the training or test dataset. The performance of the models was assessed using the missing data model weights to calculate weighted versions of accuracy, the Brier score (mean squared error), logarithmic loss, the AUC-ROC, and the area under the precision-recall curve (AUC-PR). The results are presented overall and stratified by diabetic status. We repeated this procedure (unweighted for the imputation models) for each of the missingness sensitivity analysis models described above (Statistical Modelling of Missingness), along with an additional weighted approach restricted to only patients with an admission diagnosis of sepsis.

##### Model interpretation

During the final step we refit the models on the entire dataset using the missingness weights, graphing relevant GAM non-linear effects for interpretation and comparison so LR coefficients. Additionally, as a sensitivity analysis we fit a XGBoost model using blood glucose, blood lactate and diabetic status and assess the resulting non-parametric estimate of the relationship between blood glucose and hospital mortality for various blood lactate levels by diabetic status.

#### Code availability

All analysis were performed using R version 4.2.1 [33] using ggplot2 [34] and gratia [35] for graphics, mgcv [21] and XGBoost [29] for statistical modelling, along with the packages listed previously. The project code is available at www.github.com/oizin/lactate.

## Results

### Patient characteristics

The characteristics of patients at ICU admission, over the first 24 hours of their ICU stay and at discharge are shown in Table 1. The table enables comparison of those patients who had a lactate measurement taken with those who did not – with more analysis of the impact of missingness found in Appendix A (Figure A1). Those who had a lactate measurement taken were on average more severely ill (Apache-IVa score: 68 vs 50), more likely to have an admission diagnosis of sepsis (33% vs 6%), more likely to be ventilated or receiving insulin (45% vs 24% and 38% vs 25%) and had poorer outcomes (hospital mortality of 16.2% vs 6.5%). The missing data weighting overcame these differences to a large degree, albeit with some residual differences. Comparing the weighted cohort to the overall cohort we see that the above differences in illness severity (Apache-IV score: 58 vs 57), sepsis diagnosis (17% vs 17%), ventilated (35% vs 32%), insulin (36% vs 30%), and outcomes (hospital mortality of 11.5% vs 10.3%) are greatly reduced. Our weighted analysis cohort is thus a marginally more severely ill version of our target cohort (Figure 1).

**Table 1.**
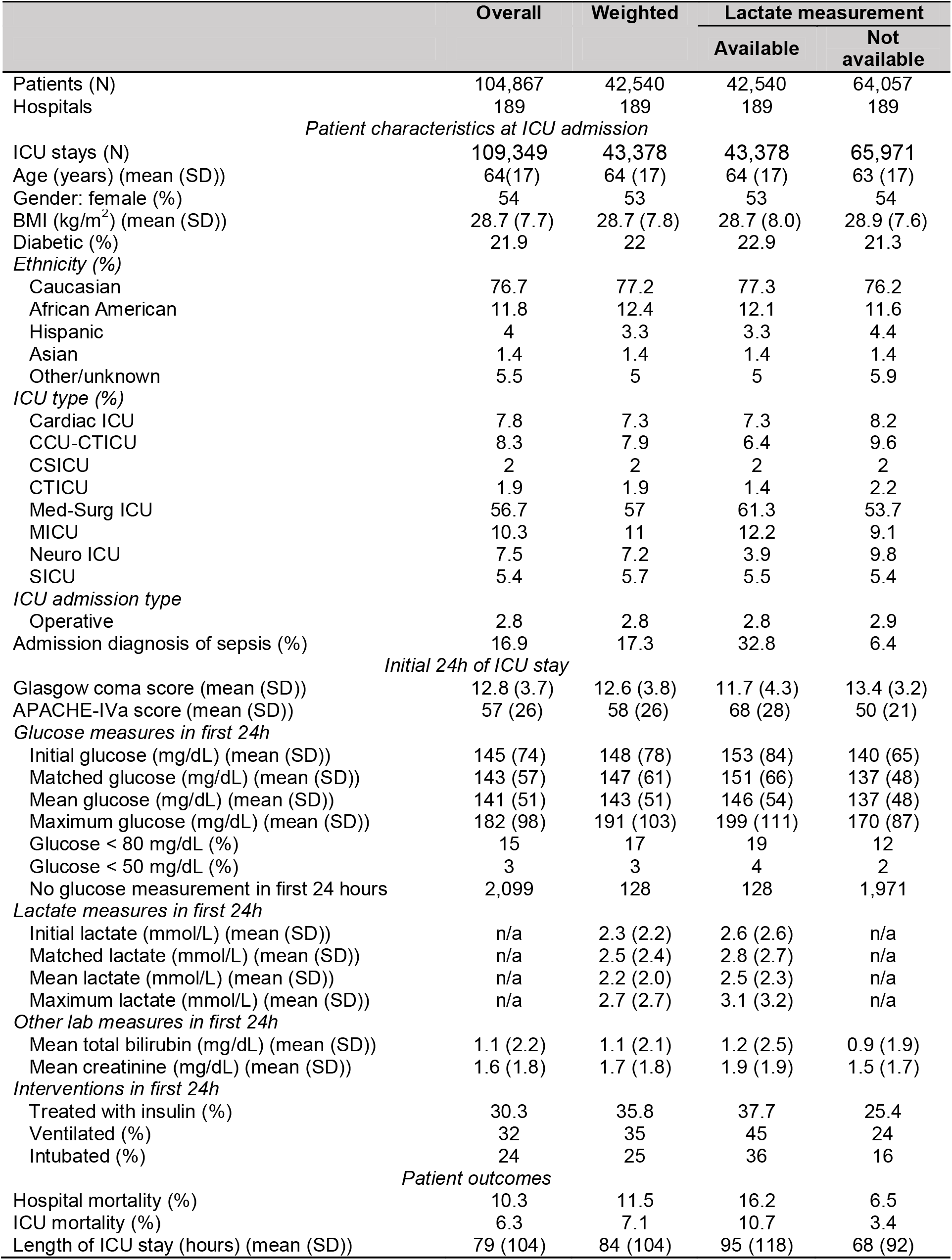
Descriptive statistics for all eligible patients (overall), the missing data weighted cohort and the overall cohort stratified by lactate availability.

The mean lactate measurement of 2.3 mmol/L in the weighted cohort compares to values of 3.8 mmol/L (hospital mortality: 20.1%) [3], 1.5 mmol/L (hospital mortality: 11.1%) [6] and 1.4 mmol/L (hospital mortality: 13.3%) [4] in previous literature. Additionally, we note that the hospital mortality rate in the 34,257 ICU stays without APACHE information was 8.7%, comparable with the target cohort with elective admissions included (hospital mortality percentage of 9.0%) and suggestive that this information could be considered missing at random.

Figure 2 outlines the univariate and bivariate relationship between blood glucose and blood lactate. As seen in Figure 2A both blood glucose and blood lactate have right skewed distributions, even after log-transformation. There is a suggestion of a U-shaped relationship between blood glucose and blood lactate, with both hyperglycaemia and hypoglycaemia associated with hyperlactatemia. Stratifying by diabetic status the results are in line with expectation (Figure 2B). Patients with diabetes have an upward shift in their blood glucose distribution. In comparison, grouping by hospital outcome reveals those who died have a greater spread in blood glucose values, in line with evidence that both hypoglycaemia and hyperglycaemia are markers of mortality risk. In contrast, the relationship for blood lactate appears simpler, with a clear upward shift in the distribution of blood lactate for those who died.

**Figure 2.**
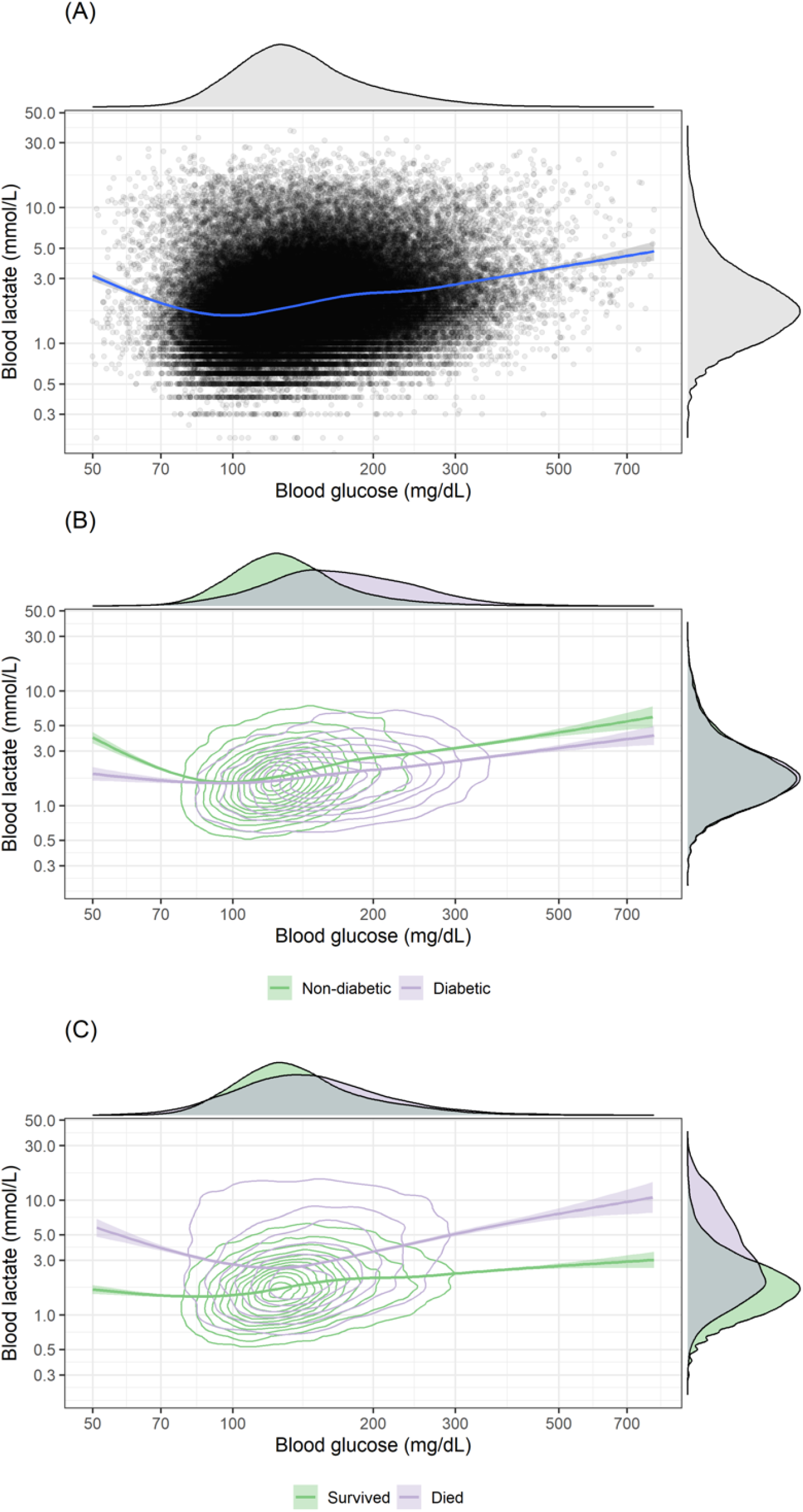
The univariate and bivariate distributions of admission blood glucose and blood lactate with fitted loess curve for: A) all data points; B) stratified by diabetic status; and C) stratified by hospital survival status.

### Missingness model

The coefficients, and measures of their uncertainty, for the variables we used to model missing lactate are shown in Table 2. As the model outcome is 1 if lactate is missing and 0 otherwise, a positive coefficient indicates increased likelihood of missingness when the variables value increases. The results are in line with Table 1, with each unit reduction in the Apache-IVa score associated with a ∼3% increase in the odds of missing lactate. Neuro ICU and operative patients are less likely to have a lactate measurement while sepsis patients are more likely. The model has good discrimination with an AUC-ROC of 0.788, illustrated in Figure 3A. Graphical analysis of model calibration (Figure 3B) reveals that calibration is generally good, with some evidence that the lower probabilities are marginal over-estimates of the chance of missingness (Figure 3A). As seen in Figure 3A the distribution of the missingness probabilities is bimodal, with a broadly sepsis group clustered around 0.25 and non-sepsis around 0.75. The resulting weights are largely less than 10, with 75% less than 4 and 50% less than 2 (Figure 3D). Results for the alternative XGBoost model and blood lactate imputation models can be found in Appendix A (Tables A2-4).

**Figure 3.**
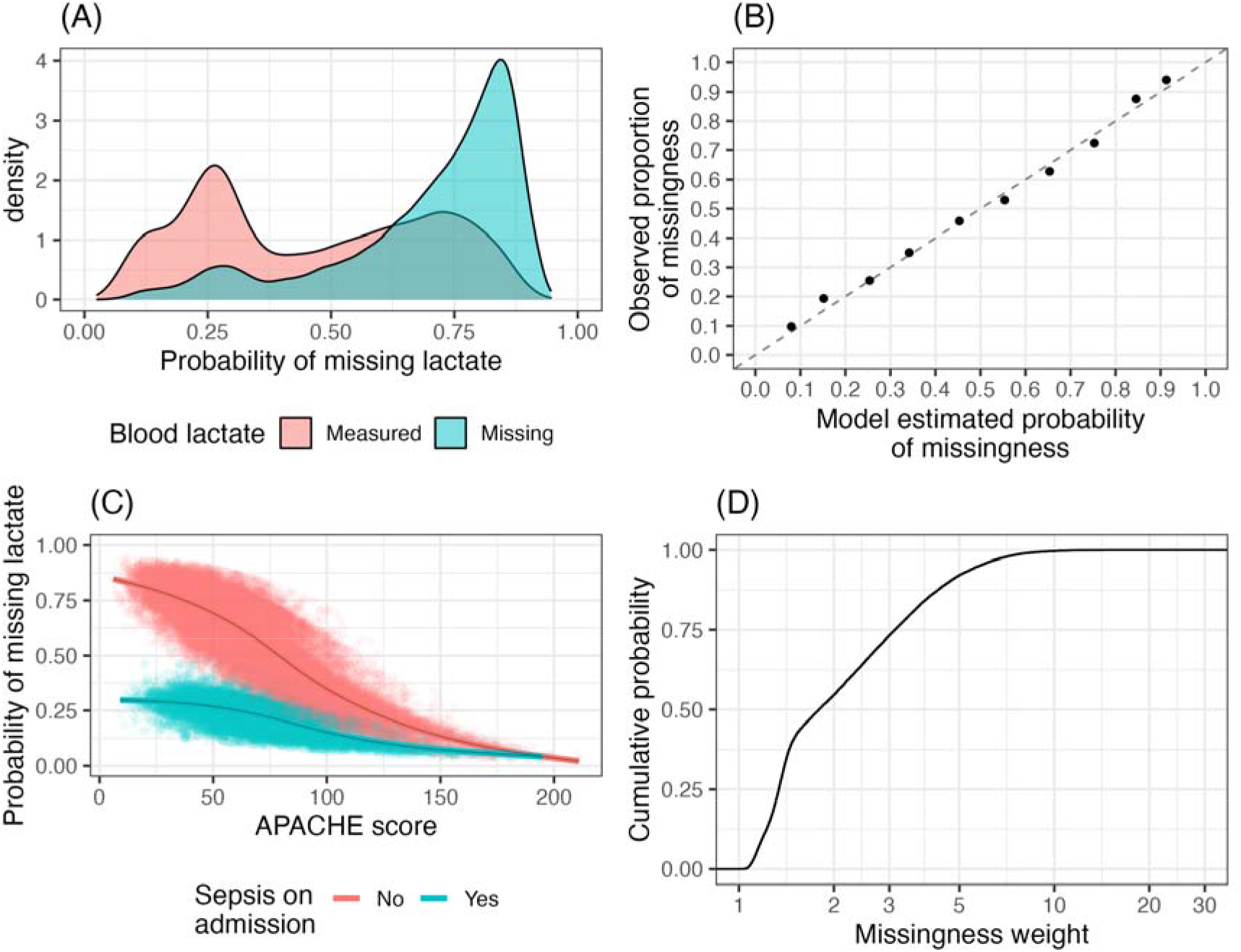
Lactate missingness model results. A) Distribution of model probability of missingness by actual missingness category, an indication of model discriminatory ability. B) Model calibration curve. Each point includes 95% CI (largely not visible due to large sample size). C) Estimated probability of missingness by APACE-IVa score and sepsis diagnosis. C) Distribution of missingness weights

**Table 2.** Missing blood lactate logistic regression model (exp. = exponentiated).

| Variable | Coefficient | Standard error | Lower 95% | Upper 95% |
| --- | --- | --- | --- | --- |
| Intercept | 1.9285 | 0.0412 | 1.8479 | 2.0092 |
| APACHE diagnosis: sepsis | -3.0040 | 0.0580 | -3.1177 | -2.8903 |
| APACHE IVa score | -0.0255 | 0.0004 | -0.0263 | -0.0247 |
| APACHE IVa score : Sepsis | 0.0145 | 0.0008 | 0.0129 | 0.0162 |
| Age | 0.0114 | 0.0005 | 0.0105 | 0.0124 |
| Operative admission | 0.1711 | 0.0428 | 0.0872 | 0.2550 |
| Ventilated | -0.2916 | 0.0255 | -0.3415 | -0.2417 |
| Intubated | -0.3252 | 0.0286 | -0.3813 | -0.2692 |
| *APACHE diagnosis organ system* |  |  |  |  |
| Gastrointestinal | -0.6809 | 0.0263 | -0.7325 | -0.6294 |
| Genitourinary | -0.9256 | 0.0444 | -1.0127 | -0.8385 |
| Hematology | -0.4828 | 0.0723 | -0.6245 | -0.3411 |
| Metabolic/Endocrine | -0.6891 | 0.0509 | -0.7888 | -0.5894 |
| Musculoskeletal/Skin | -0.8738 | 0.0872 | -1.0447 | -0.7030 |
| Neurologic | -0.0371 | 0.0240 | -0.0842 | 0.0100 |
| Respiratory | -0.5382 | 0.0224 | -0.5821 | -0.4943 |
| Transplant | -1.2388 | 0.4179 | -2.0578 | -0.4198 |
| Trauma | -0.3687 | 0.0380 | -0.4433 | -0.2942 |
| *ICU type* |  |  |  |  |
| CCU-CTICU | 0.1979 | 0.0364 | 0.1266 | 0.2692 |
| CSICU | -0.1922 | 0.0562 | -0.3024 | -0.0820 |
| CTICU | 0.1269 | 0.0596 | 0.0100 | 0.2438 |
| Med-Surg ICU | -0.0522 | 0.0274 | -0.1059 | 0.0014 |
| MICU | -0.0203 | 0.0336 | -0.0861 | 0.0455 |
| Neuro ICU | 0.5352 | 0.0411 | 0.4547 | 0.6157 |
| SICU | -0.0546 | 0.0398 | -0.1326 | 0.0234 |

### Model selection

The results of the model cross-validation are given in Table 3 and show variation in performance across the models compared to the reference model *GAM: lactate*.

**Table 3.** Weighted cross-validation results for all patients and stratified by diabetic status.

| Model | AUC-ROC | AUC-PR | Log loss | Brier |
| --- | --- | --- | --- | --- |
| *Overall (N=43,250)* |  |  |  |  |
| *LR: glucose* | 0.558 | 0.137 | 0.355 | 0.102 |
| *LR: lactate* | 0.631 | 0.164 | 0.346 | 0.100 |
| *LR: glucose + lactate* | 0.638 | 0.171 | 0.345 | 0.099 |
| *LR: glucose + lactate + (glucose : insulin)* | 0.640 | 0.173 | 0.345 | 0.099 |
| *GAM: glucose* | 0.577 | 0.153 | 0.354 | 0.101 |
| *GAM: lactate* | 0.659 | 0.232 | 0.338 | **0.097** |
| *GAM: glucose + lactate* | 0.664 | 0.239 | 0.337 | **0.097** |
| *GAM: (glucose : insulin)* | **0.665** | **0.244** | **0.336** | **0.097** |
| *APACHE-IVa* | 0.823 | 0.412 | 0.263 | 0.076 |
| *Non-diabetic (N=33,310)* |  |  |  |  |
| *LR: glucose* | 0.577 | 0.153 | 0.357 | 0.103 |
| *LR: lactate* | 0.636 | 0.169 | 0.349 | 0.101 |
| *LR: glucose + lactate* | 0.650 | 0.184 | 0.347 | 0.101 |
| *LR: glucose + lactate + (glucose : insulin)* | 0.651 | 0.185 | 0.347 | 0.101 |
| *GAM: glucose* | 0.598 | 0.173 | 0.355 | 0.102 |
| *GAM: lactate* | 0.663 | 0.235 | 0.342 | 0.099 |
| *GAM: glucose + lactate* | **0.675** | **0.252** | **0.339** | **0.098** |
| *GAM: (glucose : insulin)* | 0.673 | 0.249 | **0.339** | **0.098** |
| *APACHE-IVa* | 0.803 | 0.369 | 0.265 | 0.075 |
| *Diabetic (N=9,940)* |  |  |  |  |
| *LR: glucose* | 0.503 | 0.106 | 0.342 | 0.096 |
| *LR: lactate* | 0.614 | 0.147 | 0.334 | 0.095 |
| *LR: glucose + lactate* | 0.620 | 0.153 | 0.334 | 0.094 |
| *LR: glucose + lactate + (glucose : insulin)* | 0.614 | 0.151 | 0.334 | 0.095 |
| *GAM: glucose* | 0.529 | 0.126 | 0.341 | 0.096 |
| *GAM: lactate* | **0.644** | **0.231** | **0.325** | **0.092** |
| *GAM: glucose + lactate* | 0.643 | **0.231** | 0.324 | **0.092** |
| *GAM: (glucose : insulin)* | **0.644** | **0.232** | **0.325** | **0.092** |
| *APACHE-IVa* | 0.829 | 0.426 | 0.262 | 0.077 |

Overall, the addition of blood glucose into the predictive models improve performance over lactate alone, with the GAMs that include blood glucose outperforming the reference model. Indeed, the addition of diabetic status and indication that insulin therapy was commenced appears to lead to further slight performance improvements. Further, the models illustrate that binning of covariates results in an inferior performance compared to the comparable GAM. As shown in Appendix A (Figure A2-6) these results were consistent across sensitivity analyses.

### Model interpretation

Based on the results of the cross-validation we refit the GAM models on the data due to their similar performance. The regression spline effects are shown in Figure 4, with the effect of blood glucose on mortality varying depending on which factors are adjusted for in the model. Adjustment for lactate alone results in a moderate attenuation of the association between hyperglycaemia (although not hypoglycaemia) and hospital outcome (Figure 4A). Further adjustment for diabetic status reveals different risk profiles for diabetics and non-diabetics (Figure 4B), with the effect of hyperglycaemia much reduced for diabetics. There is evidence of an interaction effect between glucose and lactate in non-diabetics with both hypo- and hyper-glycaemia associated with a poorer outcome compared to normo-glycaemia for a given blood lactate level (Figure 4C). This interaction effect is attenuated for diabetics (Figure 4D). These results are largely consistent with the XGBoost based sensitivity analysis reported in Appendix A (Figure A8) which suggests an interaction effect whereby the risk profile for blood glucose is less attenuated at higher blood lactate values.

**Figure 4.**
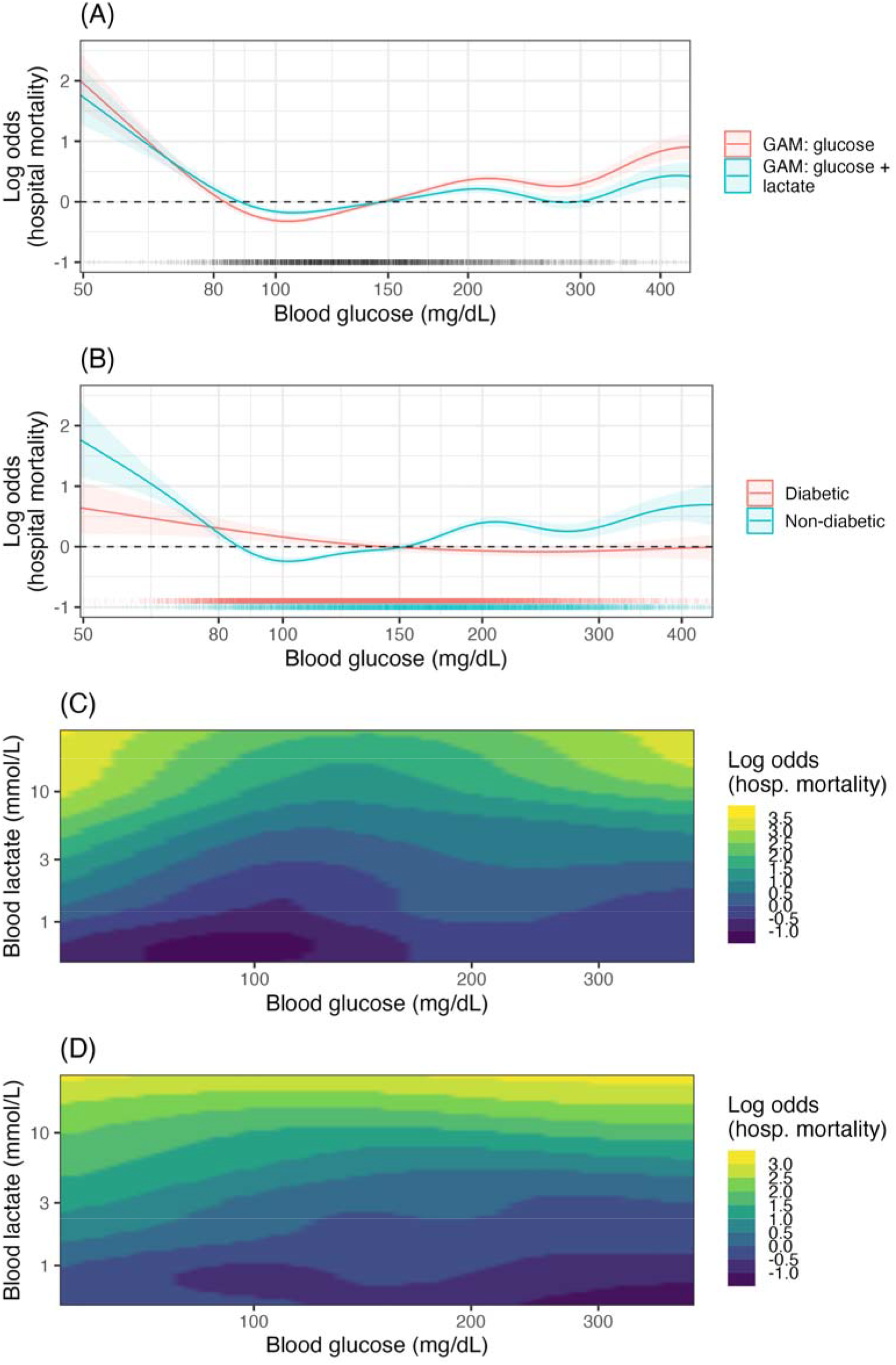
GAM model partial effects (log odds scale) for several GAM models. A) The impact of adjustment for lactate on the partial effect of glucose. B) The partial effects of glucose for diabetics and non-diabetics accounting for insulin treatment *GAM: glucose | (DM : insulin) + lactate.* C) The 2D spline interaction effect between blood glucose and lactate levels for non-diabetics *GAM: (glucose : lactate | DM = 0).* D) The 2D spline interaction effect between blood glucose and lactate levels for diabetics *GAM: (glucose : lactate | DM = 1)*.

## Discussion

The aim of this research project was to investigate the relationship between blood glucose and hospital mortality while accounting for blood lactate measurements. Across 43,250 ICU admissions, weighted to account for missing data, we assessed the predictive ability of several models stratified by diabetic status. Additionally, we varied the functional form of the model, using binning or flexible semi-parametric GAMs to model the continuous variables. We found that inclusion of blood glucose improved predictive performance. Assessment of the functional form of the GAMs revealed that in non-diabetics hyperglycaemia remained a risk factor for hospital mortality with a lessened effect in diabetics. In both subgroups hypoglycaemia remained a risk. Sensitivity analyses using alternative models and approaches to accounting for missing data supported these findings.

### Relationship to previous literature

The relationship between hyperglycaemia, hyperlactatemia and outcome in critically ill patients has been assessed in previous research. Two early studies found conflicting results on the impact of adjusting for lactate on the association between glucose and hospital outcome. Martin, Blobner [5] found both glucose and lactate on admission to be independent predictors of mortality in a study of 1,551 surgical ICU patients, with an 18 mg/dL increase in blood glucose associated with a 1% increase in the odds of mortality when controlling for lactate. In contrast, Kaukonen, Bailey [4] found that inclusion of lactate nullified the predictive power of glucose on hospital outcome in 7,925 mixed ICU patients. A common feature of these studies is the use of glucose as linear term in logistic regression. Later studies using discretisation – whether through stratification or binning – found an interactive effect between the two variables. Freire Jorge, Wieringa [6] found similar results to the present research, with abnormal levels of both blood glucose and blood lactate associated with the poorest outcomes, a finding we reproduced (see Figure 4C and LR coefficient table in Appendix A Table A5). Additionally in the current study, using two-dimensional spline terms, we found an interaction effect such that with increased blood lactate the lowest risk blood glucose level increased, from approximately 100 mg/dL to 150 mg/dL as blood lactate rose from 1 to 10 mmol/L (Figure 4C). In an alternative approach Chen, Bi [3] assessed the additional impact of lactate on glucose’s prognostic value finding it added value for patients with hypo- and hyperglycaemia. Examining the results of the current study, these findings, while superficially contradictory, may be explained by variation in study design and analysis techniques as discussed further below.

The current results are in line with previous research suggesting that admission blood glucose has a different risk profile for diabetics and non-diabetics. It has been suggested that chronic exposure to hyperglycaemia in diabetes results in metabolic adaption, reducing the potential toxic effects of acute hyperglycaemia in critical illness [36]. While no mechanism for this elevation has been described, higher HbA1c has been associated with an elevated renal threshold for glucose [37], illustrating that diabetes may result in altered physiological homeostasis, albeit with significant negative side-effects [38]. Indeed, several authors have found a blunted or absent relationship between acute hyperglycaemia and mortality risk in diabetics [39–42], in line with the current findings. On the other hand, there is evidence that the threshold for hypoglycaemic risk is elevated in those with uncontrolled diabetes [43]. In the current study we found that patients with diabetes have an inflection towards greater risk around 100-150 mg/dL (Figure 4B), higher than standard definitions of hypoglycaemia, but in line with American Diabetes Association guidelines that a range of >140 mg/dL is appropriate for hospitalised diabetics [44].

### Implications of study findings

Two features of the previous research on glucose and lactate stand out in relationship to the current study. None of the previous studies reported results either stratified by diabetic status or equivalently adjusted using interaction terms. As discussed above and shown in the current study (Figure 4B), the relationship between blood glucose and mortality is modified by diabetic status. Given the “diabetic pandemic”, adjusting for diabetic status (or HbA1c level) when investigating the relationship between blood glucose and an outcome is clearly important.

In situations where risk profiles (or other physiological effects) may be non-linear, or interactive, model choice is important, and differences may explain variation in the previous results. As suggested by Freire Jorge, Wieringa [6] the use of glucose as linear term may partially explain the results in Kaukonen, Bailey [4]. More generally, we found that the binned models underperformed the spline-based GAM models despite no loss in model interpretability (Table 3). At the extreme, choice of bin cut point to account for non-linear effects can lead to spurious results [45]. While purely non-linear methods such as deep learning are increasingly used to account for non-linear effects, such models remain largely uninterpretable, with interpretability metrics not even guaranteed to give similar findings [46]. Thus these models, as currently implemented, are not ideal for informing high stakes decisions such as those made in medicine [47], or for answering certain scientific questions. GAMs and other “white-box” machine learning approaches [48] present an alternative approach for situations where non-linear effects are expected and interpretability is key.

### Strengths and weaknesses

This paper has several strength and weakness. Strengths noted above are comparison of several model choices and the inclusion of mediating treatment factors such as insulin in the models. The predictive aspect of model selection enables easy comparison of models with complex interactive effects and different estimation methods [49]. Through varying the model form and assessing predictive ability overall and across subgroups we gain a greater idea of the true discriminatory power of the models in the studied population. Other strengths include the use of a large high quality real world data collection [22, 23]. The data source consists of a mixed ICU population across a wide geographic area, with potential variation in treatment practises and cohorts treated. However, the retrospective observational nature of the data source is also a weakness. The data were not originally designed for use in this study, and a significant portion of patients in the source database did not have a blood lactate measurement. Missing data is a known cause of bias in analysis of EMR data, with sicker patients typically have more complete records, as seen in the current findings [50, 51]. While inverse probability of missingness weighting offers a straightforward approach to designing analyses aimed at reducing this bias the weighting did not completely remove differences between the groups (Table 1), and fundamentally cannot account for unmeasured predictors of missingness.

### Future directions

While this research has confirmed that blood glucose is a marker of poor prognosis even in the presence of stronger markers (blood lactate) further research is required into the potential mechanisms of any causal effect on patient outcome along with subgroup variation. Ultimately lactate is produced from glucose, and discounting the presence of tissue hypoxia [12], both hyperglycaemia and hyperlactatemia are measures of altered energy metabolism. However, biophysiological theories of how acute stress disturbances in energy metabolism cause poor outcomes are only starting to be pieced together [7, 14]. Given the long timeframes of RCTs [52] future research should continue aiming to link theory and observational data. For instance, if as claimed by Gunst, De Bruyn [15] the treatment effect of insulin is entirely through reducing glucose toxicity, it should be possible to demonstrate this in observational data using methods to estimate causal effects of time-varying exposures that treat blood glucose measures as mediators such as g-computation [53]. Previous research looking at the treatment effect of insulin in observational data has found a negative impact of insulin treatment [54], largely explained by huge differences in rates of hypoglycaemia (29% vs. 1.4%) between treated and non-treated. Causal inference in observational data is notoriously difficult and a wider range of study designs need to be applied to the data to investigate this issue, in particular looking at subgroup effects (e.g. diabetic vs. non-diabetic or surgical vs. non-surgical patients [15]).

## Conclusion

In a mixed ICU population admission blood glucose is predictive of hospital mortality after accounting for blood lactate, with different hyperglycaemic risk profiles for diabetics and non-diabetics. In diabetics we found no association between hyperglycaemia and hospital mortality, while in non-diabetics hyperglycaemia remains a predictor of poor outcomes.

## Appendix A: Additional results

### Descriptive statistics

**Figure A1.**
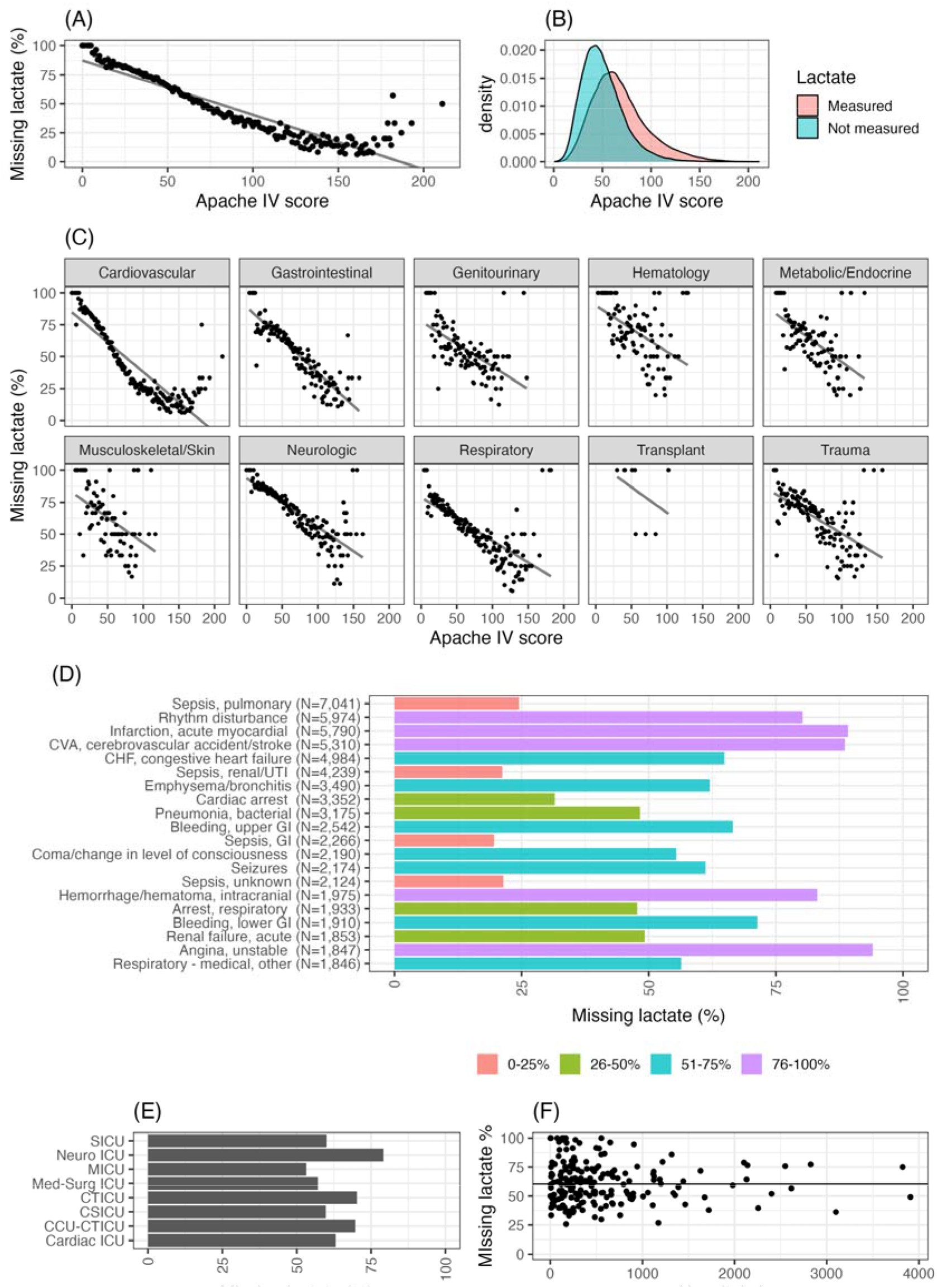
Characterisation of ICU stays with and without a blood lactate measurement. A) APACHE-IVa score and percentage of missing lactate. B) Distribution of APACHE-IVa score by indication of missing lactate. C) APACHE-IVa score and percentage of missing lactate by admission organ system. D) Top 20 admission diagnoses and percentage of missing lactate. E) ICU type and percentage of missing lactate. F) Hospital size and percentage of missing lactate.

**Table A1.** Alternative version of Table 1 using the XGBoost weights.

|  | Overall | Weighted | Lactate measurement |  |
| --- | --- | --- | --- | --- |
|  |  |  | Available | Not available |
| Patients (N) | 104,867 | 42,540 | 42,540 | 64,057 |
| Hospitals | 189 | 189 | 189 | 189 |
| *Patient characteristics at ICU admission* |  |  |  |  |
| ICU stays (N) | 109,349 | 43,378 | 43,378 | 65,971 |
| Age (years) (mean (SD)) | 64(17) | 63 (17) | 64 (17) | 63 (17) |
| Gender: female (%) | 54 | 53 | 53 | 54 |
| BMI (kg/m<sup>2</sup>) (mean (SD)) | 28.7 (7.7) | 28.7 (7.8) | 28.7 (8.0) | 28.9 (7.6) |
| Diabetic (%) | 21.9 | 22 | 22.9 | 21.3 |
| *Ethnicity (%)* |  |  |  |  |
| Caucasian | 76.7 | 76.9 | 77.3 | 76.2 |
| African American | 11.8 | 12.3 | 12.1 | 11.6 |
| Hispanic | 4.0 | 3.2 | 3.3 | 4.4 |
| Asian | 1.4 | 1.4 | 1.4 | 1.4 |
| Other/unknown | 5.5 | 5.5 | 5 | 5.9 |
| *ICU type (%)* |  |  |  |  |
| Cardiac ICU | 7.8 | 7.0 | 7.3 | 8.2 |
| CCU-CTICU | 8.3 | 8.4 | 6.4 | 9.6 |
| CSICU | 2.0 | 2.4 | 2.0 | 2.0 |
| CTICU | 1.9 | 2.3 | 1.4 | 2.2 |
| Med-Surg ICU | 56.7 | 56.1 | 61.3 | 53.7 |
| MICU | 10.3 | 10.7 | 12.2 | 9.1 |
| Neuro ICU | 7.5 | 7.7 | 3.9 | 9.8 |
| SICU | 5.4 | 5.5 | 5.5 | 5.4 |
| *ICU admission type* |  |  |  |  |
| Operative | 2.8 | 2.8 | 2.8 | 2.9 |
| Admission diagnosis of sepsis (%) |  |  |  |  |
| *Initial 24h of ICU stay* |  |  |  |  |
| Glasgow coma score (mean (SD)) | 12.8 (3.7) | 12.7 (3.7) | 11.7 (4.3) | 13.4 (3.2) |
| APACHE-IVa score (mean (SD)) | 57 (26) | 57 (26) | 68 (28) | 50 (21) |
| *Glucose measures in first 24h* |  |  |  |  |
| Initial glucose (mg/dL) (mean (SD)) | 145 (74) | 147 (75) | 153 (84) | 140 (65) |
| Matched glucose (mg/dL) (mean (SD)) | 143 (57) | 146 (59) | 151 (66) | 137 (48) |
| Mean glucose (mg/dL) (mean (SD)) | 141 (51) | 142 (50) | 146 (54) | 137 (48) |
| Maximum glucose (mg/dL) (mean (SD)) | 182 (98) | 187 (100) | 199 (111) | 170 (87) |
| Glucose < 80 mg/dL (%) | 15 | 16 | 19 | 12 |
| Glucose < 50 mg/dL (%) | 3 | 3 | 4 | 2 |
| No glucose measurement in first 24 hours | 2,099 | 128 | 128 | 1,971 |
| *Lactate measures in first 24h* |  |  |  |  |
| Initial lactate (mmol/L) (mean (SD)) | n/a | 2.27 (2.22) | 2.55 (2.58) | n/a |
| Matched lactate (mmol/L) (mean (SD)) | n/a | 2.49 (2.41) | 2.80 (2.73) | n/a |
| Mean lactate (mmol/L) (mean (SD)) | n/a | 2.21 (1.96) | 2.49 (2.32) | n/a |
| Maximum lactate (mmol/L) (mean (SD)) | n/a | 2.68 (2.70) | 3.08 (3.15) | n/a |
| *Other lab measures in first 24h* |  |  |  |  |
| Mean total bilirubin (mg/dL) (mean (SD)) | 1.06 (2.21) | 1.03 (2.06) | 1.22 (2.49) | 0.92 (1.91) |
| Mean creatinine (mg/dL) (mean (SD)) | 1.62 (1.78) | 1.63 (1.82) | 1.85 (1.89) | 1.46 (1.67) |
| *Interventions in first 24h* |  |  |  |  |
| Treated with insulin (%) | 30.3 | 34.5 | 37.7 | 25.4 |
| Ventilated (%) | 32 | 33 | 45 | 24 |
| Intubated (%) | 24 | 24 | 36 | 16 |
| *Patient outcomes* |  |  |  |  |
| Hospital mortality (%) | 10.3 | 11.0 | 16.2 | 6.5 |
| ICU mortality (%) | 6.3 | 6.8 | 10.7 | 3.4 |
| Length of ICU stay (hours) (mean (SD)) | 79 (104) | 82 (102) | 95 (118) | 68 (92) |

### Missingness and imputation models

#### Missingness models

The XGBoost missingness model (Table A2) has an AUC-ROC of 0.823.

**Table A2.** Variable importance (top 20) for the XGBoost missingness model

| Feature | Gain | Gain SD |
| --- | --- | --- |
| Sepsis | 0.2308 | 0.0018 |
| Apache IVa score | 0.1279 | 0.0113 |
| Bicarbonate | 0.1062 | 0.0021 |
| Intubated | 0.0570 | 0.0120 |
| Calcium | 0.0537 | 0.0015 |
| Chloride | 0.0452 | 0.0024 |
| Potassium | 0.0381 | 0.0013 |
| BMI | 0.0386 | 0.0009 |
| Sodium | 0.0388 | 0.0013 |
| Creatinine | 0.0331 | 0.0016 |
| BUN | 0.0305 | 0.0017 |
| Age | 0.0252 | 0.0006 |
| Ventilated | 0.0242 | 0.0034 |
| CVA | 0.0150 | 0.0005 |
| MI | 0.0145 | 0.0006 |
| GCS | 0.0122 | 0.0022 |
| Pneumonia | 0.0076 | 0.0004 |
| Neuro ICU | 0.0079 | 0.0007 |
| Unstable angina | 0.0062 | 0.0004 |

#### Imputation models

The linear regression (GAM) blood lactate imputation model (Table A3) had a RMSE of 2.513, while the XGBoost model (Table A4) had a RMSE of 2.412 mmol/L.

**Table A3.**
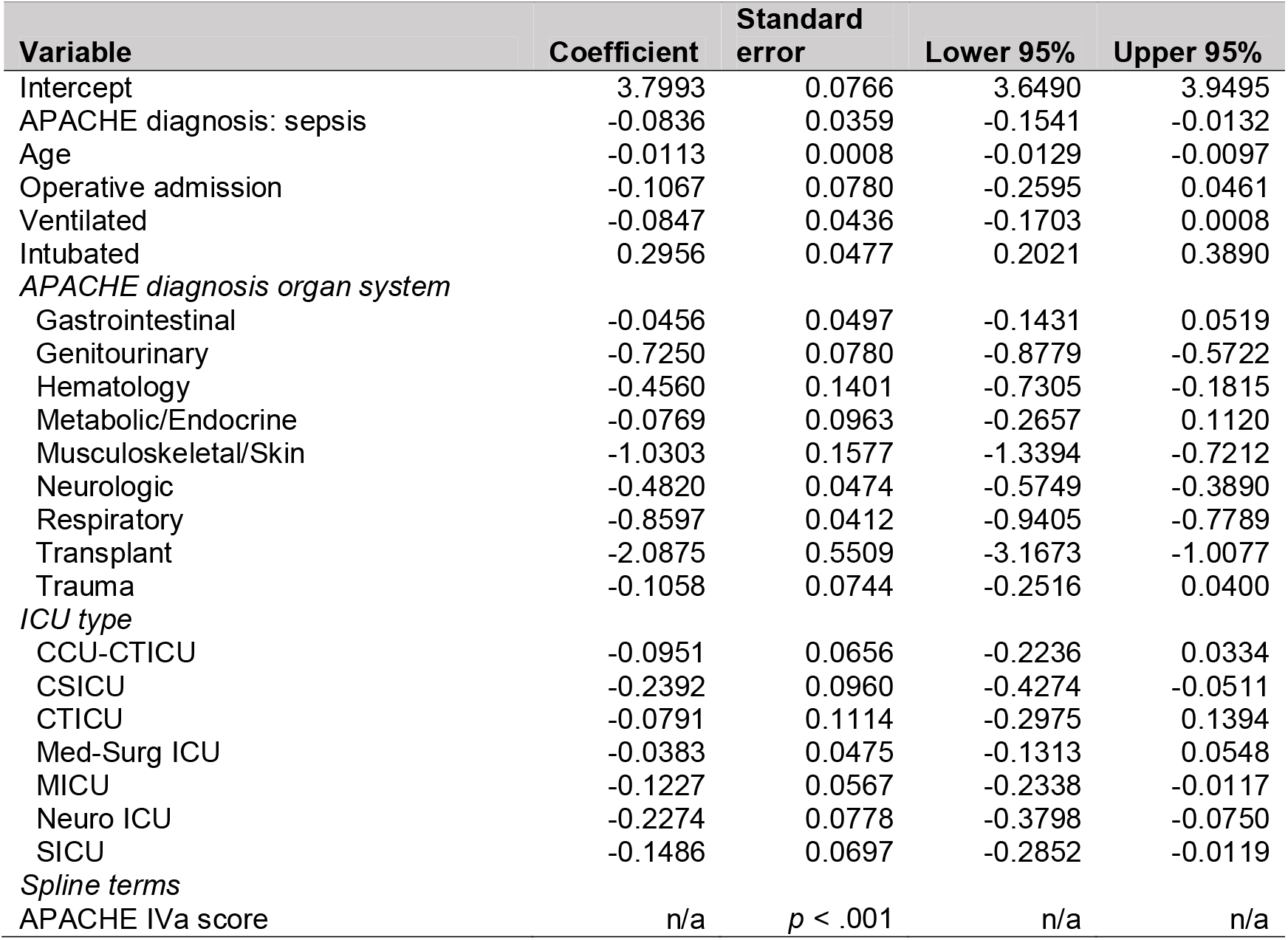
Blood lactate linear GAM model

| Variable | Coefficient | Standard error | Lower 95% | Upper 95% |
| --- | --- | --- | --- | --- |
| Intercept | 3.7993 | 0.0766 | 3.6490 | 3.9495 |
| APACHE diagnosis: sepsis | -0.0836 | 0.0359 | -0.1541 | -0.0132 |
| Age | -0.0113 | 0.0008 | -0.0129 | -0.0097 |
| Operative admission | -0.1067 | 0.0780 | -0.2595 | 0.0461 |
| Ventilated | -0.0847 | 0.0436 | -0.1703 | 0.0008 |
| Intubated | 0.2956 | 0.0477 | 0.2021 | 0.3890 |
| *APACHE diagnosis organ system* |  |  |  |  |
| Gastrointestinal | -0.0456 | 0.0497 | -0.1431 | 0.0519 |
| Genitourinary | -0.7250 | 0.0780 | -0.8779 | -0.5722 |
| Hematology | -0.4560 | 0.1401 | -0.7305 | -0.1815 |
| Metabolic/Endocrine | -0.0769 | 0.0963 | -0.2657 | 0.1120 |
| Musculoskeletal/Skin | -1.0303 | 0.1577 | -1.3394 | -0.7212 |
| Neurologic | -0.4820 | 0.0474 | -0.5749 | -0.3890 |
| Respiratory | -0.8597 | 0.0412 | -0.9405 | -0.7789 |
| Transplant | -2.0875 | 0.5509 | -3.1673 | -1.0077 |
| Trauma | -0.1058 | 0.0744 | -0.2516 | 0.0400 |
| *ICU type* |  |  |  |  |
| CCU-CTICU | -0.0951 | 0.0656 | -0.2236 | 0.0334 |
| CSICU | -0.2392 | 0.0960 | -0.4274 | -0.0511 |
| CTICU | -0.0791 | 0.1114 | -0.2975 | 0.1394 |
| Med-Surg ICU | -0.0383 | 0.0475 | -0.1313 | 0.0548 |
| MICU | -0.1227 | 0.0567 | -0.2338 | -0.0117 |
| Neuro ICU | -0.2274 | 0.0778 | -0.3798 | -0.0750 |
| SICU | -0.1486 | 0.0697 | -0.2852 | -0.0119 |
| *Spline terms* |  |  |  |  |
| APACHE IVa score | n/a | $p < .001$ | n/a | n/a |

**Table A4.** Variable importance (top 20) for the XGBoost imputation model

| Feature | Gain | Gain SD |
| --- | --- | --- |
| Bicarbonate | 0.2451 | 0.0067 |
| Apache IVa score | 0.2352 | 0.0050 |
| Chloride | 0.1220 | 0.0066 |
| Sodium | 0.1018 | 0.0068 |
| BUN | 0.0567 | 0.0029 |
| APACHE diagnosis: Cardiac Arrest | 0.0519 | 0.0033 |
| Creatinine | 0.0458 | 0.0039 |
| Potassium | 0.0216 | 0.0014 |
| Calcium | 0.0214 | 0.0033 |
| BMI | 0.0178 | 0.0011 |
| Age | 0.0146 | 0.0018 |
| APACHE diagnosis: Seizure | 0.0111 | 0.0018 |
| Intubated | 0.0074 | 0.0015 |
| APACHE diagnosis: Sepsis | 0.0068 | 0.0006 |
| APACHE diagnosis: Upper GI bleed | 0.0048 | 0.0012 |
| GCS (verbal) | 0.0038 | 0.0005 |
| GCS | 0.0037 | 0.0009 |
| oobventday1 | 0.0018 | 0.0013 |
| APACHE diagnosis: GI bleed | 0.0012 | 0.0004 |

### Model selection

The following tables (A2-6) graph the cross-validation results across various sensitivity analysis except Table A3 which repeats the results in Table 3.

**Figure A2.**
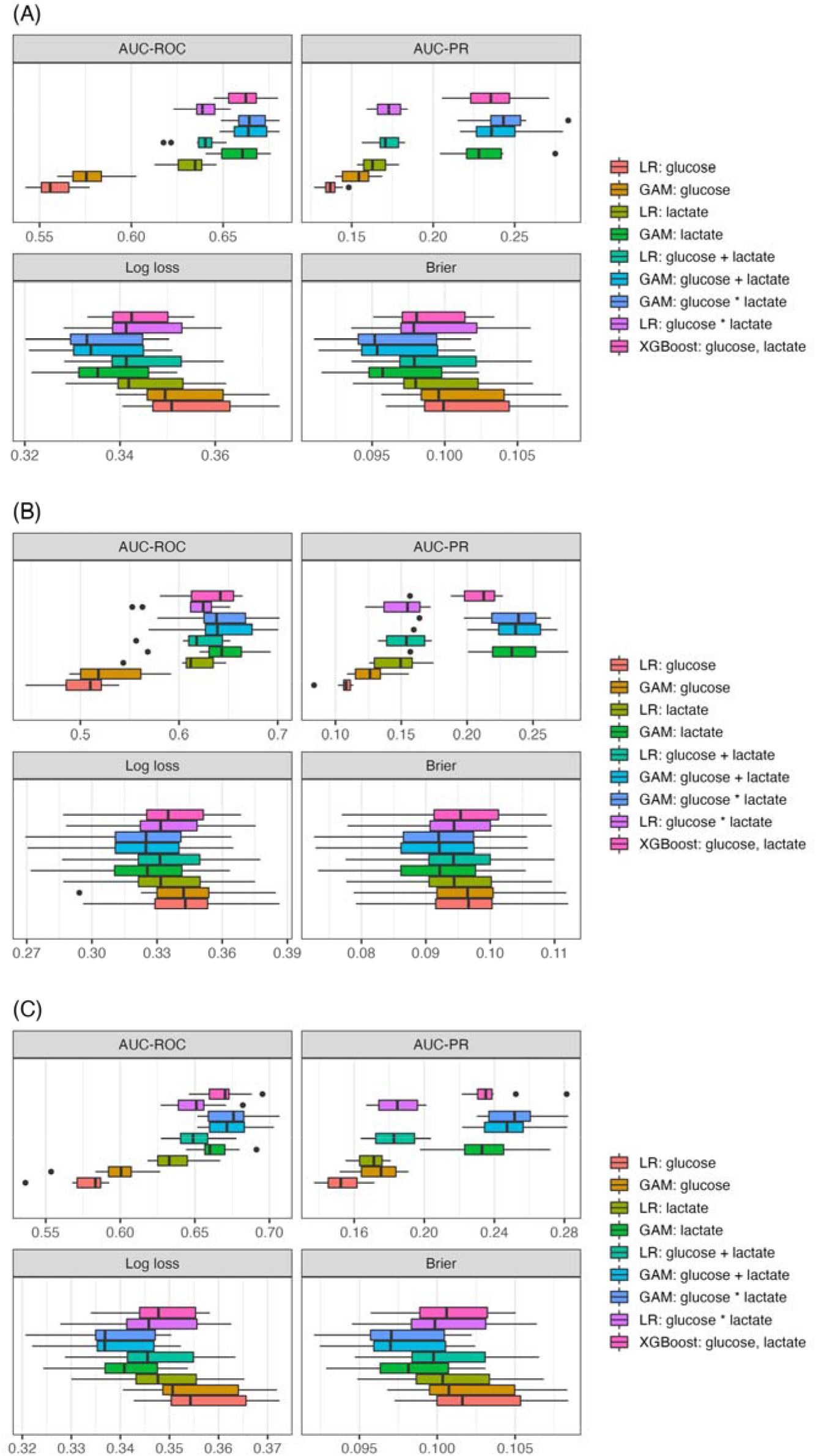
Weighted cross-validation results using logistic regression generated weights (A) All patients (B) Diabetics. (C) Non-diabetics.

**Figure A3.**
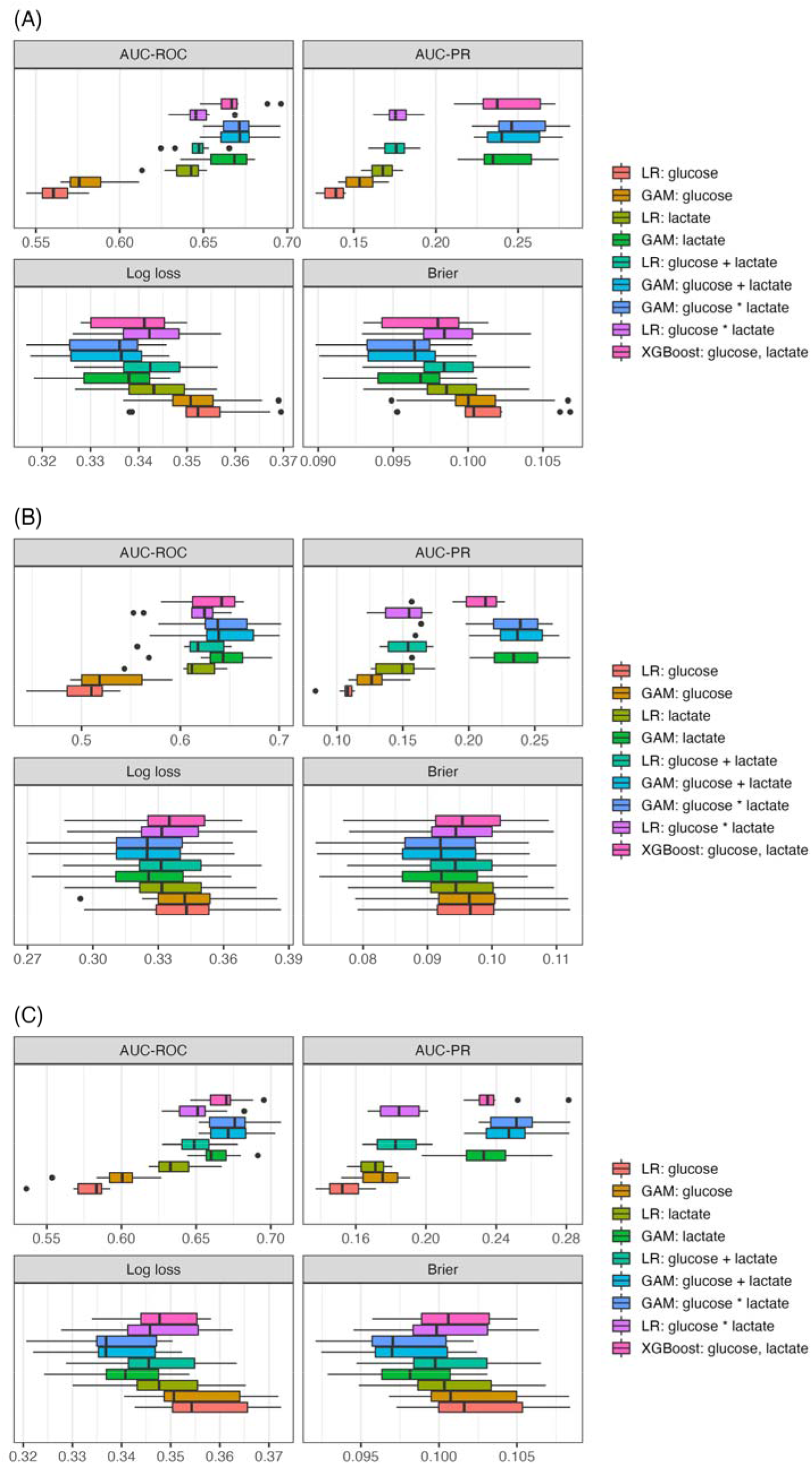
Weighted cross-validation results using XGBoost generated weights (A) All patients (B) Diabetics. (C) Non-diabetics.

**Figure A4.**
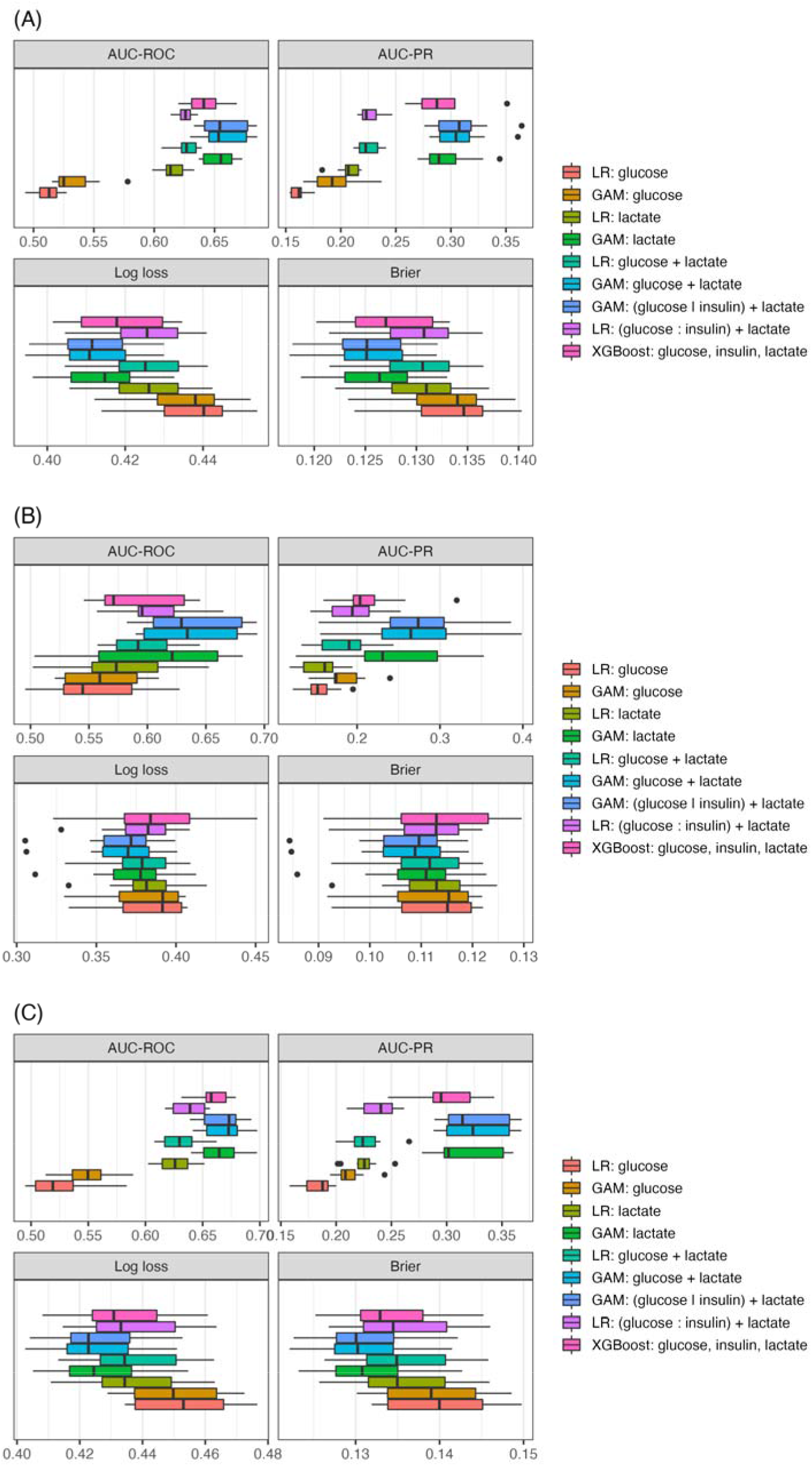
Weighted cross-validation results for patients with an admission diagnosis of Sepsis using averaged weights (A) All patients (B) Diabetics. (C) Non-diabetics.

**Figure A5.**
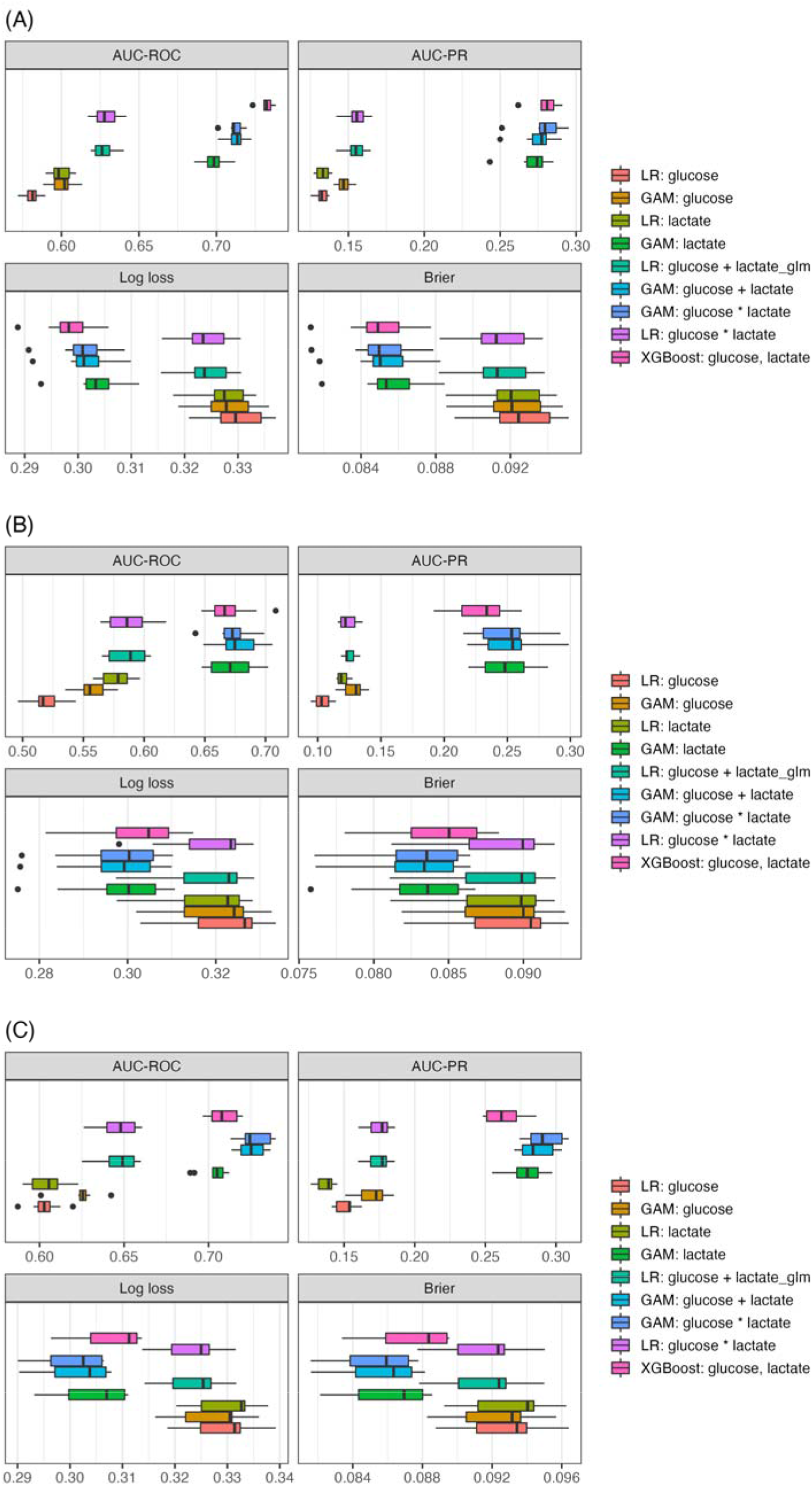
Cross-validation results with missing lactate values imputed using the linear

**Figure A6.**
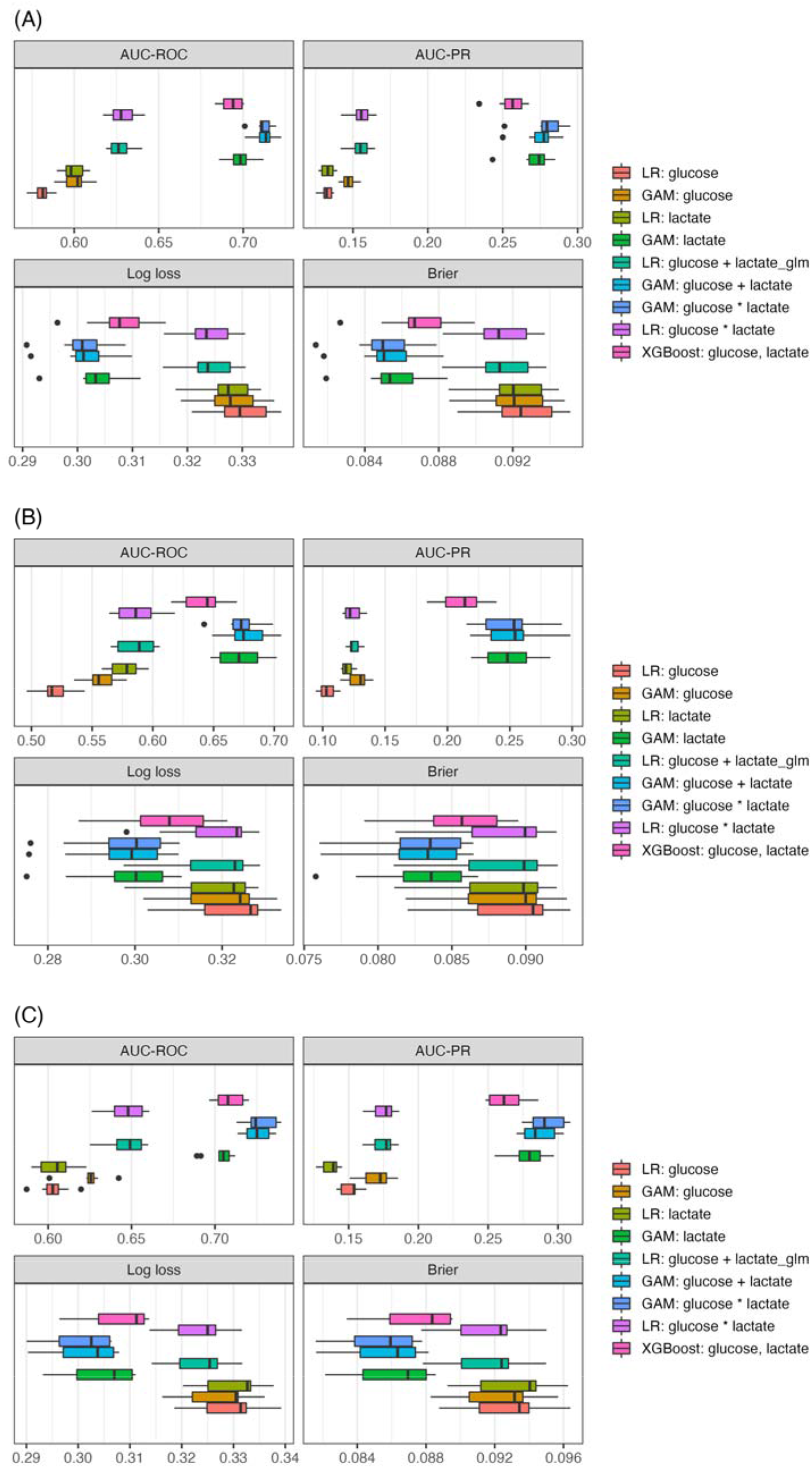
Cross-validation results with missing lactate values imputed using the

### Model interpretation

The following tables (A7-8) graph the impact of blood glucose on hospital mortality.

**Figure A7.**
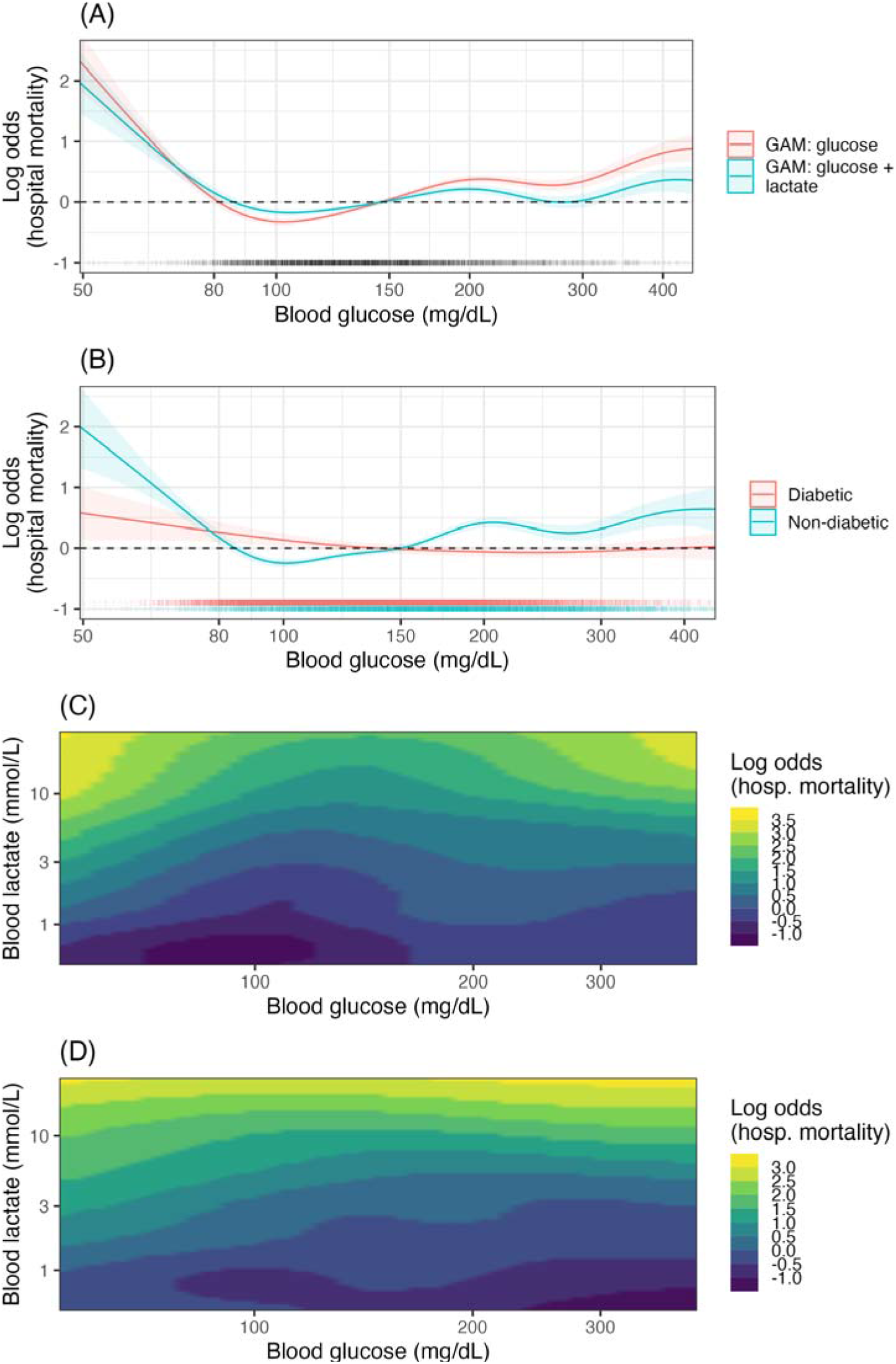
GAM model partial effects (log odds scale) for several GAM models using the XGBoost generated weights. A) The impact of adjustment for lactate on the partial effect of glucose. B) The partial effects of glucose for diabetics and non-diabetics accounting for insulin treatment *GAM: glucose | (DM : insulin) + lactate.* C) The 2D spline interaction effect between blood glucose and lactate levels for non-diabetics *GAM: (glucose : lactate | DM = 0).* D) The 2D spline interaction effect between blood glucose and lactate levels for diabetics *GAM: (glucose : lactate | DM = 1)*.

**Figure A8.**
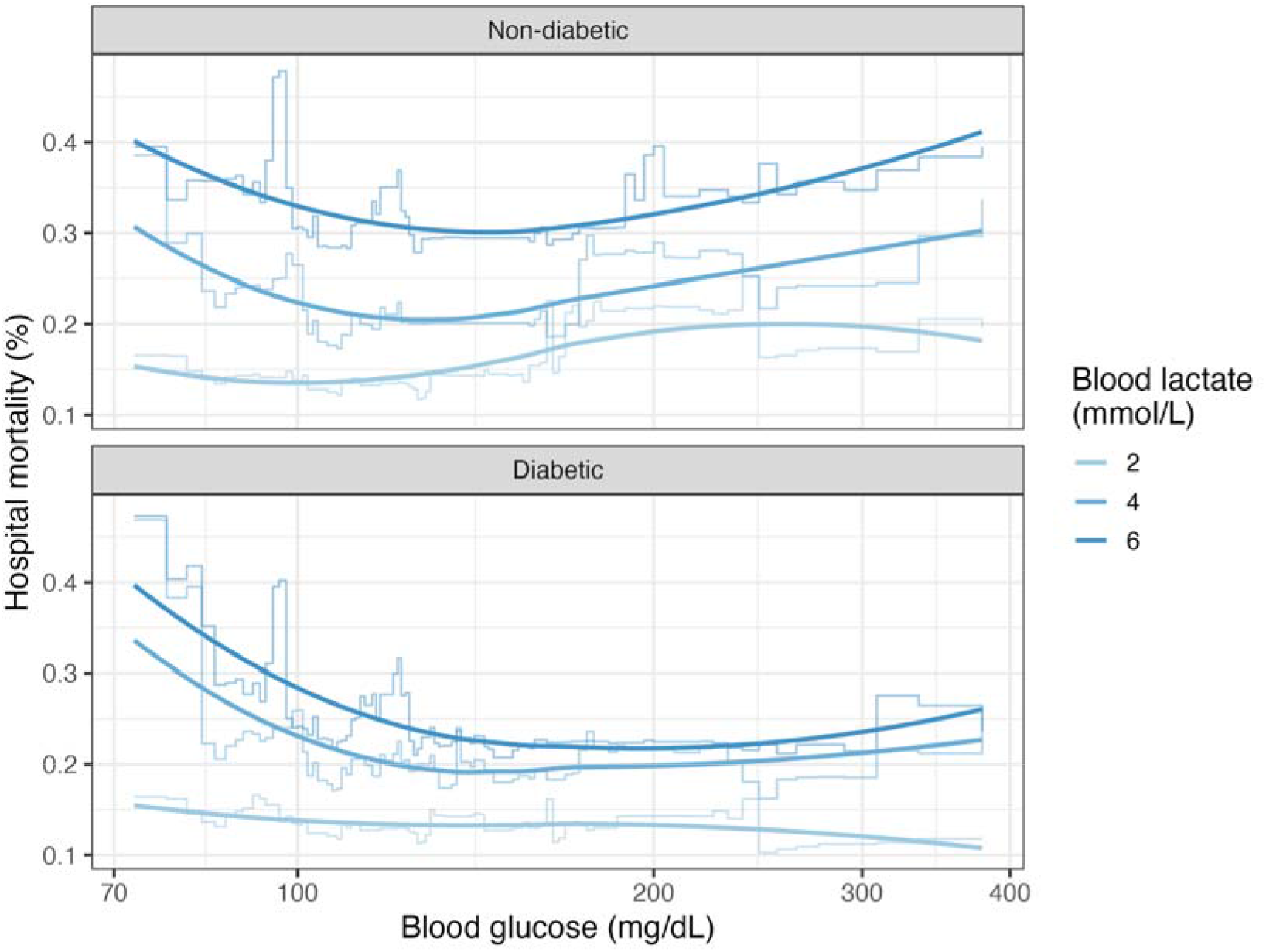
Nonparametric conditional estimates of mortality risk by blood glucose and blood lactate levels stratified by diabetic status using XGBoost

**Table A5.** Logistic regression model with hospital mortality as outcome and blood glucose and blood lactate interaction terms

| Variable | Coefficient | Standard error | Statistic | p value |
| --- | --- | --- | --- | --- |
| (Intercept) | -3.081 | 0.064 | -48.026 | 0.000 |
| Blood glucose (126-137] | 0.307 | 0.139 | 2.217 | 0.027 |
| Blood glucose (137-148] | 0.359 | 0.147 | 2.446 | 0.014 |
| Blood glucose (148-162] | 0.430 | 0.165 | 2.602 | 0.009 |
| Blood glucose (162+) | 0.727 | 0.125 | 5.792 | 0.000 |
| Blood lactate (1-1.3] | 0.438 | 0.082 | 5.312 | 0.000 |
| Blood lactate (1.3-1.7] | 0.543 | 0.085 | 6.351 | 0.000 |
| Blood lactate (1.7-2.3] | 0.656 | 0.079 | 8.330 | 0.000 |
| Blood lactate (2.3+) | 1.442 | 0.064 | 22.635 | 0.000 |
| Blood glucose (126-137]:Blood lactate (1-1.3] | -0.374 | 0.205 | -1.824 | 0.068 |
| Blood glucose (137-148]:Blood lactate (1-1.3] | 0.078 | 0.201 | 0.385 | 0.700 |
| Blood glucose (148-162]:Blood lactate (1-1.3] | 0.058 | 0.222 | 0.264 | 0.792 |
| Blood glucose (162+):Blood lactate (1-1.3] | -0.141 | 0.174 | -0.811 | 0.418 |
| Blood glucose (126-137]:Blood lactate (1.3-1.7] | -0.108 | 0.203 | -0.532 | 0.595 |
| Blood glucose (137-148]:Blood lactate (1.3-1.7] | -0.126 | 0.216 | -0.583 | 0.560 |
| Blood glucose (148-162]:Blood lactate (1.3-1.7] | -0.266 | 0.232 | -1.143 | 0.253 |
| Blood glucose (162+):Blood lactate (1.3-1.7] | -0.194 | 0.170 | -1.140 | 0.254 |
| Blood glucose (126-137]:Blood lactate (1.7-2.3] | -0.257 | 0.186 | -1.386 | 0.166 |
| Blood glucose (137-148]:Blood lactate (1.7-2.3] | -0.184 | 0.197 | -0.934 | 0.350 |
| Blood glucose (148-162]:Blood lactate (1.7-2.3] | -0.441 | 0.214 | -2.066 | 0.039 |
| Blood glucose (162+):Blood lactate (1.7-2.3] | -0.065 | 0.152 | -0.430 | 0.667 |
| Blood glucose (126-137]:Blood lactate (2.3+) | -0.395 | 0.156 | -2.531 | 0.011 |
| Blood glucose (137-148]:Blood lactate (2.3+) | -0.493 | 0.165 | -2.986 | 0.003 |
| Blood glucose (148-162]:Blood lactate (2.3+) | -0.505 | 0.179 | -2.818 | 0.005 |
| Blood glucose (162+):Blood lactate (2.3+) | -0.391 | 0.133 | -2.926 | 0.003 |

